# Human In the Loop Challenges for Quality Annotation of Pre-Cancer Lesions in Clinical Oral Images

**DOI:** 10.64898/2026.07.02.26355859

**Authors:** Sourav Mandal, Pramila Mendonca, Keerthi Gurusanth, Harshita Thakur, Praveen Birur, Anupama Shetty, Debnath Pal

## Abstract

Smartphone-based Artificial Intelligence (AI) enabled screening of the hyperplasia and dysplasia stages (pre-cancer, OPMD) offers a viable opportunity to reduce the incidence and mortality of oral cancer through early prevention. However, developing accurate segmentation models requires high-quality, pixel-level annotations that are prohibitively expensive and prone to clinical subjectivity. To address this, we empirically validated a deep learning-driven Human-in-the-Loop (HITL) iterative pseudo-labeling framework to pixel-annotate 3,026 clinical oral images. We also conducted controlled experiments to quantify the network’s tolerance to label noise (unreviewed pseudo-labels) and resolved clinical subjectivity using pixel-wise Cohen’s Kappa and the STAPLE consensus algorithm. While iterative self-training consistently improved lesion detection and spatial localization, including even a modest fraction (*→* 10%) of unreviewed pseudo-labels increased training convergence instability three-to-four-fold and induced a conservative prediction bias that negatively impacted model recall. Ultimately, the model’s performance converged with the inter-rater reliability ceiling (*κ →* 0.65) against a multi-expert ground truth, successfully falling within the envelope of human agreement. These findings highlight that non-experts can successfully drive the early-to-mid stages of the annotation pipeline, but a final expert-driven quality assurance step is strictly essential to mitigate training instability, confirmation bias, and clinically unacceptable recall drops. Overall, this provides a scalable, empirically validated blueprint for building domain-specific medical imaging datasets in global health settings where annotation cost, inter-observer variability, and expert unavailability challenges are most acute.

## 1 Introduction

Oral cancer (OC) ranks among the top three cancers by incidence and mortality in India, contributing nearly a third of the global burden [1–3]. The primary driver of high mortality is delay in diagnosis due to inadequate healthcare access; consequently, 60–80% of patients in India are diagnosed in advanced, often incurable stages, compared to roughly 40% in developed nations [1]. Early screening of Oral Potentially Malignant Disorders (OPMDs), commonly called oral pre-cancer, is paramount for reducing the incidence through early intervention. Since the clinical manifestations of OC and OPMD as oral cavity lesions are well-understood and accessible for visual inspection, they present a unique opportunity for early intervention, especially in low resource settings. With low-skilled workforce, adjuncts such as Artificial Intelligence (AI)-assisted screening of oral lesions using mobile smartphones based on white light imaging provide a robust mitigation opportunity. It presents a highly scalable and cost-effective option for deployment in resource-constrained settings that need improvement across diverse applications [4].

A key requirement for a highly accurate AI classification model is to consistently focus attention on the correct regions of the image containing the relevant lesion(s). This ability to directly dissect the region of interest (ROI) / lesion(s) [5] through image segmentation provides an alternative way to understand the true attention computed by the AI model. When AI classification model attends to diverse image data during learning, it is often confused about the patterns to memorize, no matter whether domain adaptation or other learning hacks are tried. In these cases, providing accurately segmented images with the correct regions of interest marked (lesions in this case) in this diverse dataset can better guide the model to the correct regions of the image for highly accurate classification. However, accessing such data for learning, also known as pixel-level annotated data in the clinical/medical domain, is costly and tedious to prepare, as human experts must review and curate it for accuracy.

Delineating or localizing the ROI is the crux of not only cancerous lesion detection, but in any general image-driven automated analysis workflow across domains - known as “image segmentation”, that produces pixel-level annotation or segmentation mask [6]. Manual marking of oral lesions is prohibitively expensive, time-consuming, and highly dependent on domain-specific clinical expertise. Furthermore, images captured in point-of-care (POC) settings with low-end smartphones introduce severe domain-specific challenges, including wide variability in ambient lighting, diverse camera hardware, motion blur, and specular reflections [7]. Consequently, early-stage OC and OPMDs often mimic benign mucosal conditions, making visual differentiation highly subjective even among specialists and leading to significant inter-observer variability. Together, obtaining segmentation mask(s) represent the primary bottleneck in achieving accurate medical AI.

Although several datasets of oral cavity images are available, they have limitations that hinder the development of low-resource POC-deployable screening tools. Many public datasets are derived from Western populations [8], which can induce severe demographic bias and domain shift when models are applied to the Indian subcontinent [9]. Other efforts have focused on specialized modalities—such as hyperspectral imaging [10, 11], autofluorescence [12, 13], or endoscopy [14]—which are largely incompatible with the low-cost smartphone screening strategy. Furthermore, the majority of recent smartphone-based image datasets rely on imprecise image-level labels and bounding boxes rather than granular polygon masks [15–17], utilize high-end proprietary hardware [10], or suffer from limited dataset sizes due to the unscalability of purely manual annotation [18–20].

To overcome the dual challenges of annotation scalability and inter-observer subjectivity, the “Human-in-the-Loop” (HITL) paradigm has emerged as a powerful hybrid methodology [21, 22]. By intelligently combining human clinical expertise with iterative Machine Learning (ML)-driven pseudo-labeling, HITL mitigates the core weaknesses of both approaches: the unscalability of pure manual annotation and the unreliability (e.g., hallucinations and confirmation bias) of pure ML systems [23, 24]. While HITL has shown great success in accelerating annotation in histopathology [21, 22, 24, 25] and guiding classification attention maps [26], its systematic application for generating granular, pixel-level instance segmentation masks for oral oncology has not been extensively explored or empirically validated.

In this study, we present a comprehensive, systematically validated Deep Learning (DL) driven HITL framework designed to annotate a large-scale oral cancer segmentation dataset using low-end smartphone images from the Indian population. Rather than simply applying AI to pre-existing data, we focus on the algorithmic and clinical dynamics of the data annotation pipeline itself. The specific contributions of this work are threefold:

1. **Iterative HITL Pipeline Validation:** We design and empirically evaluate a multi-stage HITL segmentation pipeline, demonstrating how iterative AI pseudo-labeling - guided by minimal clinical intervention, can progressively scale dataset size while actively improving model boundary precision and spatial localization over successive cycles.
2. **Analysis of Label Noise and Convergence Dynamics:** We provide a controlled empirical analysis of the network’s tolerance to intermediate, unreviewed pseudo-labels (label noise), quantifying its impact on training instability, convergence dynamics, and precision-recall trade-offs.
3. **Probabilistic Resolution of Clinical Subjectivity:** We curate a robust dataset of 3,026 pixel-level annotated images and introduce a structured methodology for resolving multi-expert disagreement. By computing pixel-wise Cohen’s Kappa to establish an inter-rater reliability ceiling, and utilizing the STAPLE (Simultaneous Truth and Performance Level Estimation) consensus algorithm [27], we ensure probabilistic, bias-mitigated ground truth (GT) boundaries for highly ambiguous lesions.

By rigorously characterizing the interactions between DL pseudo-labeling, label noise, and human expert consensus, this work establishes a highly efficient, scalable blueprint for developing pixel-level annotated data for domain-specific medical AI models in resource-constrained global health settings.

## 2 Results

The entire workflow from the initial images and expert labels to the 3026 annotated images is summarized, and the work stages are quantitatively dissected in Fig. 1, and Fig. 2.

**Figure 1:**
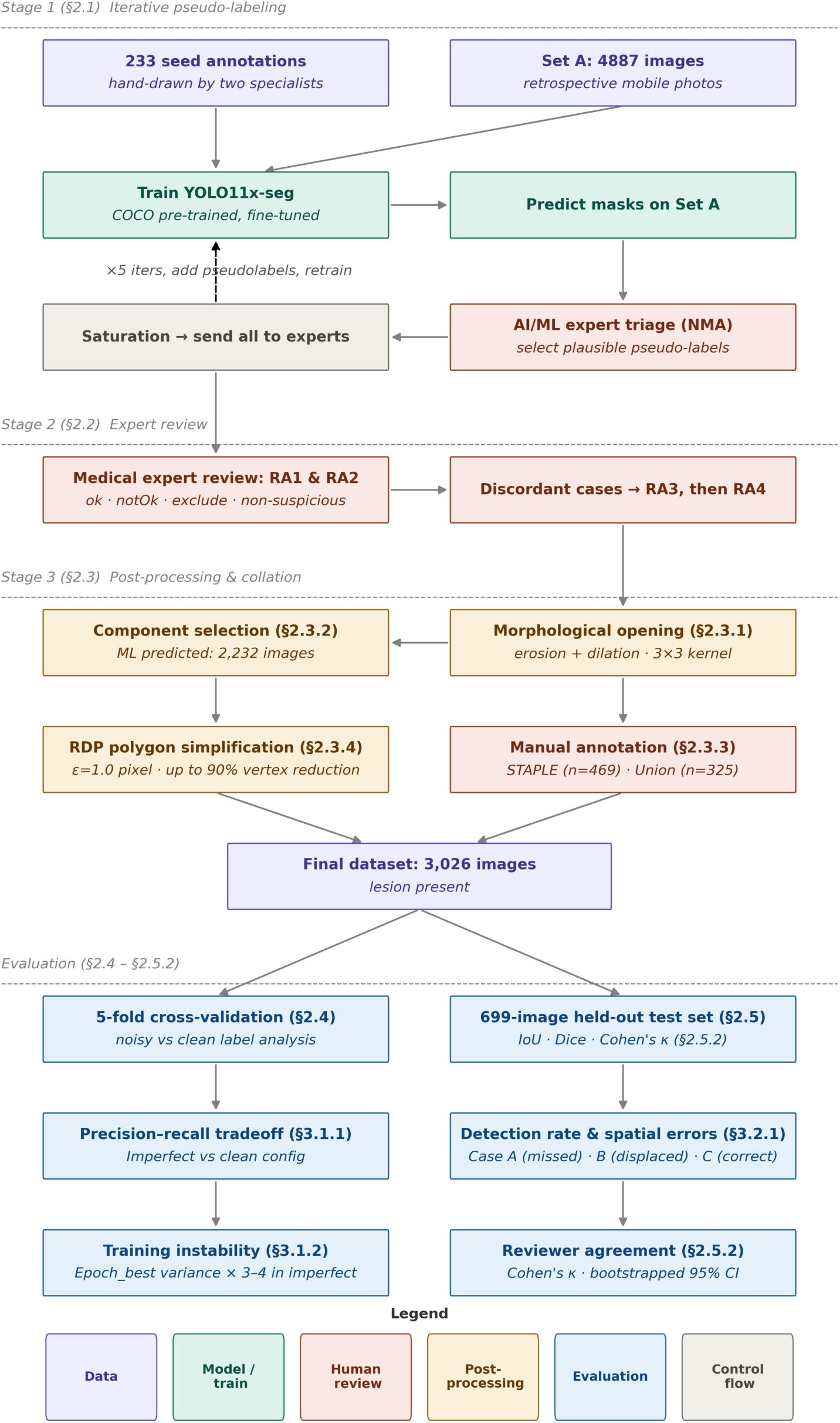
An overview of the entire study design, involving HITL annotation, label (mask), post-processing

**Figure 2:**
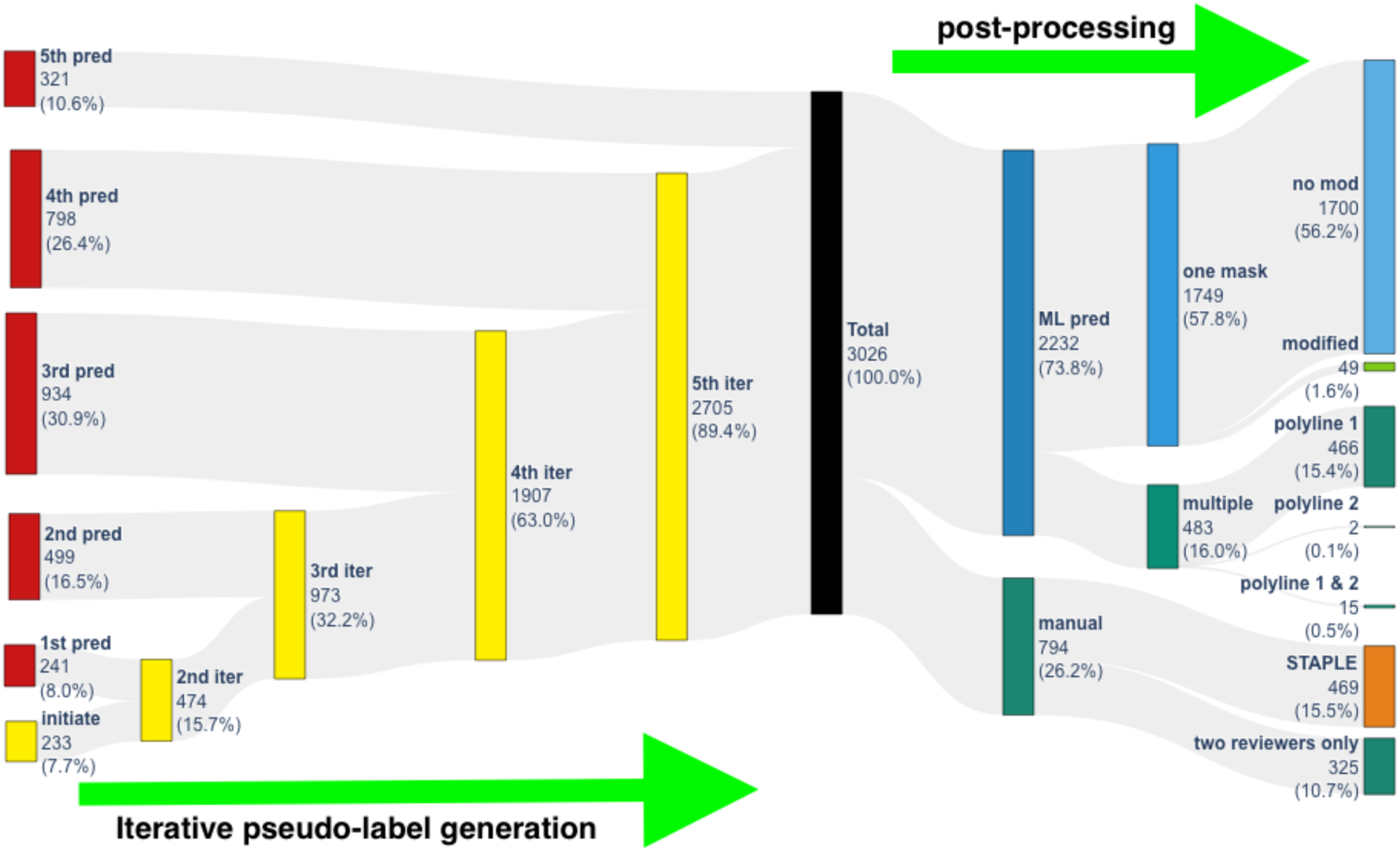
Diagram showing the different stages of HITL image annotation based on our study design outlined in Fig. 1. The number of images associated with each stage of annotation is indicated beside each bar. The stages where an iterative pseudo-label generation was deployed are indicated with an arrow at the bottom of the figure. The different post-processing methods applied to the different types and numbers of generated lesion masks are indicated under the arrow at the top right of the figure.

In the first few cycles of the iterative pseudo-labeling, the models predicted lower-quality annotations; these were included in the training, nevertheless, to prevent stagnation at the initial stage, as the volume of annotation predictions was relatively low. These images, along with others exhibiting annotation quality issues, e.g. Fig. 3a showing the original image, and Fig. 3b overlaid with the DL-predicted “unclean” mask. Such images were manually reviewed and sorted out by the NMA (non-medical annotator), and underwent morphological image processing (as described in section 2.1) to retain the largest area (Fig. 3c, blue area), and were then subjected to a final manual inspection of all annotations. We also see an example where the model failed to generate any prediction and Reviewer-cum-Annotator (RA)s had to manually draw the ROI mask (Fig. 3d).

**Figure 3:**
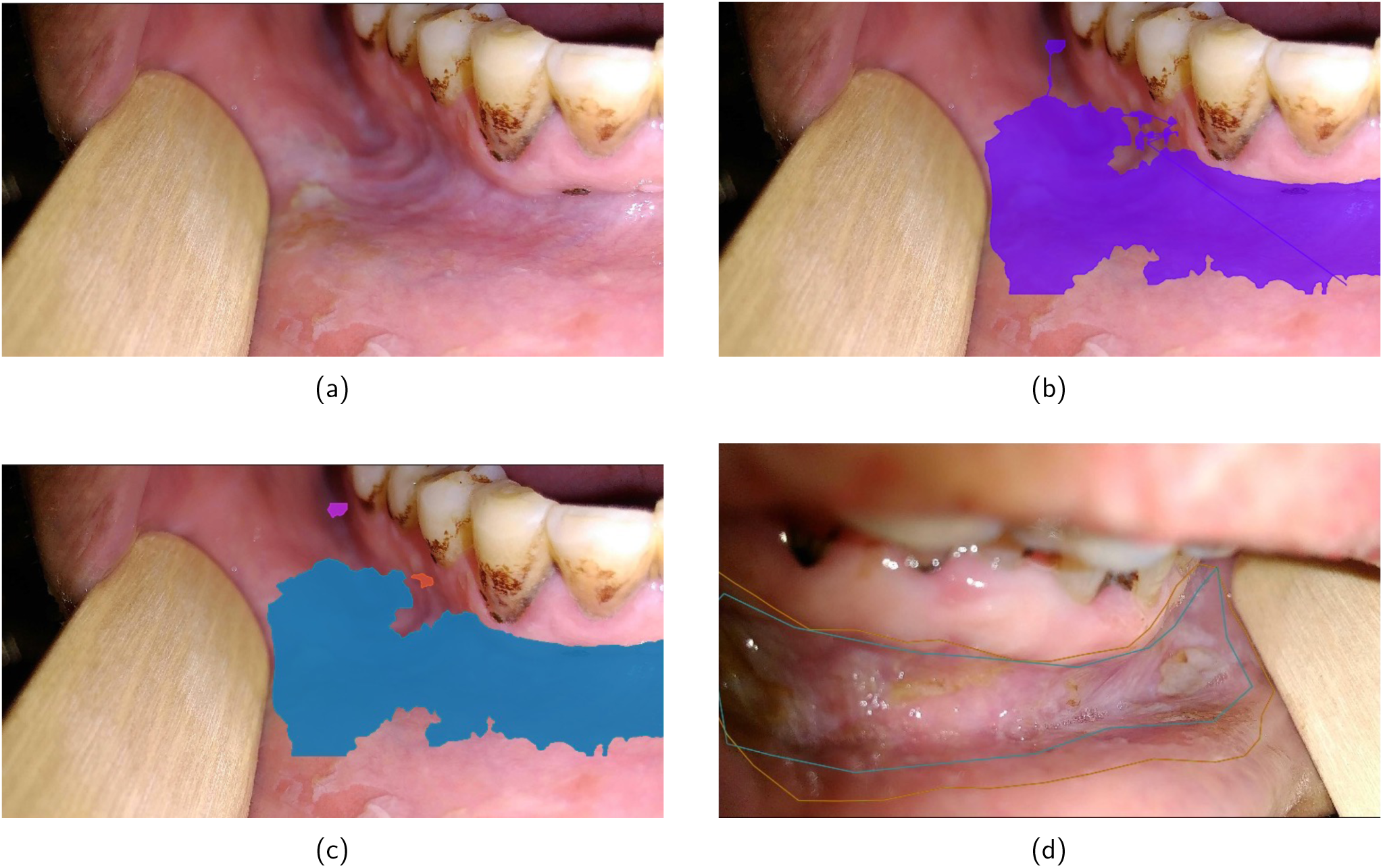
Example outputs from the HITL pipeline for pseudo labeling. (a) Original Image. (b) DL-predicted with cuts and small stray regions. (c) Morphological processing for improving mask quality by polygon separation and extraction. (d) An image where the model failed to predict the proper ROI for the lesion. Two of the reviewers’ manual annotations are depicted by a line boundary.

### 2.1 Post-processing of correct pseudo-labels

The DL-based segmentation produced a single mask in the majority of cases, but not in all, as detailed in the post-processing stages (Methods Section 4.2.3). Fig. 4A shows that opening masks for the set of 2232 images created up to 12 components but the largest single (and in some cases the first two: C1 and C2) component(s) contributed most to the total mask area (*>* 98%). For this reason, we discarded all polygon mask components other than component 1 (C1, in some cases C2 was chosen), for majority of the images (1749, Fig. 2). The median largest mask area had about 0.45M pixels or covered *<* 5% of the total images area (see Fig. 4B & supplementary Fig. S1). Among this, 49 images had to be further curated by hand for quality assurance (mask overflowing on to external non-lesion areas). The remaining 1700 needed no further modification at the final quality check (Fig. 2). However, for a considerable number of images (16% of the total set), the second largest area (represented by C2) contributed up to 50% of the total mask area (refer to Figs. 2 & 4A). These 483 images and the corresponding masks were rechecked manually, and the mask components were chosen in either of three ways (Fig. 2):

- 15 images for which both the largest (C1) and second-largest (C2) components were accepted as the final mask received two polygon entries.
- 2 images had only polyline 2 (C2) as the correct acceptable mask.
- For the remaining 466, only polyline 1 (C1) was kept as the mask.

**Figure 4:**
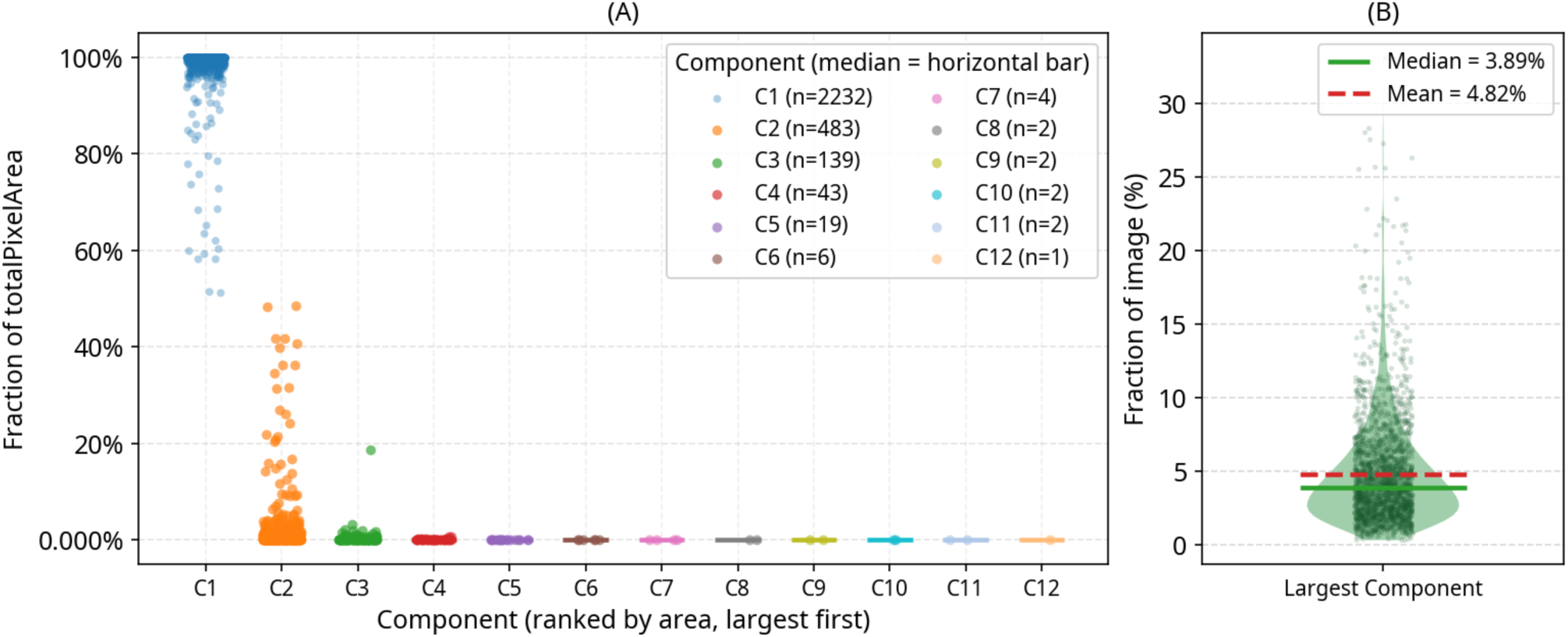
Insight into the mask components at the post-processing stage of 2232 images (ML-predicted). (A) Number of disconnected components after the opening operation and their contribution to the total mask area. (B) The area distribution of, and fraction of area covered by the largest component.

Each sample was written in two annotation formats - a LabelMe-compatible JSON with polygon vertices denormalized to absolute pixel coordinates (using per-image width and height), and a YOLO segmentation TXT with normalized coordinates prefixed by class index 0. Also, discarding sub-pixel deviations with the Ramer-Douglas-Peucker (RDP) algorithm removed digitization noise without sacrificing lesion boundary fidelity, and in practice, reduced polygon vertex counts by up to 90%, substantially decreasing annotation file sizes.

Table 1 summarizes the pseudo-labeling throughput and validation performance of the yolo11x-seg model across all five iterative HITL training cycles. The training set grew from 233 images (seed annotations) to 2,705 images by iteration 5. The detection rate on Set A, the proportion of the remaining unannotated images for which the model predicted at least one mask, peaked at 61.9% in iteration 3 and then declined in iterations 4 and 5 as the residual unannotated images increasingly comprised visually atypical cases not yet well represented in the training distribution. Mask validation mean average precision metrics, e.g. mAP50 improved substantially from 79.38 (iter. 1) to 93.07 (iter. 5), and mAP50-95 rose from 41.22 to 53.29 over the same range, indicating progressive improvement in both detection and boundary localization quality as pseudo-labels accumulated. The following subsections report the controlled experiment on label noise tolerance (Sec. 2.2) and the performance of each successive model on the fixed 699-image held-out test set (Sec. 2.3).

**Table 1:** Iterative training of the segmentation model (mask) incorporating NMA-reviewed prediction from the previous step

| Iter. | Images | Train | Val | Predicted (%) | Detection Set | Epoch<sub>best</sub> | Precision | Recall | mAP50 | mAP50-95 |
| --- | --- | --- | --- | --- | --- | --- | --- | --- | --- | --- |
| 1 | 233 | 188 | 45 | 2020 (43.40) | 4654 | 269 | 82.43 | 71.40 | 79.38 | 41.22 |
| 2 | 474 | 429 | 45 | 2411 (54.63) | 4413 | 214 | 73.29 | 78.26 | 80.28 | 38.31 |
| 3 | 973 | 928 | 45 | 2423 (61.91) | 3914 | 298 | 88.58 | 84.35 | 85.85 | 48.75 |
| 4 | 1907 | 1862 | 45 | 1278 (42.89) | 2980 | 556 | 91.30 | 84.78 | 90.96 | 53.74 |
| 5 | 2705 | 2660 | 45 | 789 (36.16) | 2182 | 277 | 92.97 | 89.13 | 93.07 | 53.29 |

### 2.2 Effect of the presence of noisy labels on the segmentation

Table 2 presents fold-level and cross-fold mean performance for both the *clean* and *imperfect* configurations across all five cross-validation folds (see Methods Section 4.2.4. Each row shows clean/imperfect values (separated by a slash); the bottom row gives the cross-fold mean and standard deviation for each condition. The following subsections examine the two principal differences observed between the two conditions: a precision-recall trade-off at the headline metric level, and a marked increase in convergence instability in the imperfect (noisy-label inclusion).

**Table 2:** Five fold cross-validation performance comparing the *Clean* dataset (pseudo-labels excluded) / *Imperfect* dataset (119 unreviewed pseudo-labels included). Rows 1-5 correspond to folds 1-5, respectively. The last two rows are averages corresponding to the *Clean* and *Imperfect* datasets, respectively.

| Epoch | Box (localization) |  |  |  | Epoch | Mask (semantic segmentation) |  |  |  |
| --- | --- | --- | --- | --- | --- | --- | --- | --- | --- |
|  | Precision | Recall | mAP50 | mAP50-95 |  | Precision | Recall | mAP50 | mAP50-95 |
| 296/534 | 87.33/88.91 | 90.28/91.24 | 93.27/94.41 | 60.24/61.03 | 340/380 | 91.31/94.77 | 91.38/87.42 | 95.11/96.57 | 62.83/65.05 |
| 369/847 | 90.82/90.51 | 90.28/91.67 | 94.67/95.17 | 60.51/60.27 | 370/494 | 89.75/91.84 | 94.79/92.36 | 96.12/95.72 | 64.06/63.45 |
| 239/185 | 88.30/95.70 | 91.75/85.08 | 94.08/95.39 | 59.82/57.90 | 351/291 | 93.86/92.65 | 92.01/89.93 | 95.27/94.82 | 62.88/62.29 |
| 251/301 | 90.01/91.88 | 90.31/88.58 | 92.80/93.51 | 58.64/57.94 | 274/418 | 88.63/89.93 | 91.75/92.04 | 94.55/94.51 | 61.70/61.91 |
| 250/288 | 89.62/91.01 | 89.28/90.75 | 93.02/94.34 | 60.22/60.22 | 293/696 | 93.66/91.48 | 88.97/88.97 | 94.97/94.65 | 63.34/62.71 |
| 281±50 | 89.22±1.36 | 90.38±0.88 | 93.57±0.74 | 59.89±0.72 | 325±38 | 91.44±2.06 | 91.78±1.88 | 95.20±0.56 | 62.96±0.80 |
| 431±243 | 91.60±2.28 | 89.46±2.52 | 94.56±0.74 | 59.47±1.34 | 455±164 | 92.13±1.73 | 90.14±2.04 | 95.25±0.78 | 63.08±1.18 |

#### 2.2.1 Precision-Recall trade-off

Despite near-identical mAP values, a consistent, directionally interpretable shift in the precision-recall balance was observed. In the imperfect condition, mask precision was higher (92.13 vs. 91.44, Δ = +0.69) while mask recall was lower (90.14 vs. 91.78, Δ = →1.64); the same pattern held for bounding box predictions (ΔPrecision = +2.38, ΔRecall = →0.92).

#### 2.2.2 Training instability and convergence dynamics

The epoch at which peak validation performance was reached (Ep_best_) exhibited dramatically higher variance in the imperfect condition. For mask training: 455 *±*164 epochs (coefficient of variation (CV) = 36%) versus 325 *±* 38 (CV = 12%); for box: 431 *±* 243 (CV = 56%) versus 281 *±* 50 (CV = 18%). Since the validation partitions differ across folds, this three-to-four-fold increase in CV is a pure training-side instability signal, not an artifact of varying validation set difficulty.

#### 2.2.3 Aggregate segmentation performance

Cross-fold mean performance metrics for both configurations are summarized in Table 2. At the headline metrics level, the two conditions performed nearly identically. Mask mAP50 was 95.25 *±* 0.78 (*imperfect*) versus 95.20 *±* 0.56 (*clean*), a difference of +0.05 percentage points, negligible relative to the cross-fold standard deviation. Mask mAP50-95, which penalizes predictions at tighter Intersection over Union (IoU) thresholds and is a more sensitive measure of boundary localization quality, differed by only +0.12 (63.08 vs. 62.96). Box mAP50 showed a slightly larger gap (94.56 vs. 93.57, Δ = +0.99), partly attributable to the larger effective training set in the imperfect condition rather than annotation quality alone. While comparing box-based learning for segmentation to mask-based learning for the same, we observe better average performance with the latter confirming that precise mask marking is an important factor in model performance.

### 2.3 Performance on an independent internal test set

#### 2.3.1 Detection rate and spatial errors

A held-out test-set of 699 images were used for evaluating the performance and learning dynamics across iterations, as described in Methods Section 4.2.5. Table 3 and Fig. 5 summarize the outcome breakdown across the five iterations (iter). Correct detection (Case C) increased from 42.1% (iter 1, 294/699 images) to 59.1% (iter 3, 413/699), declined in iter 4 (46.5%, 325/699), and recovered to 62.4% (iter 5, 436/699). The overall trend is non-monotonic but upward, with iter 5 achieving the highest detection rate of the five models (Fig. 5A, C). Spatial errors (Case B) decreased sharply from 9.9% (iter 1) to 1.6% (iters 4 and 5), indicating that later models became markedly better calibrated in their *spatial selectivity* - when they predicted, they increasingly did so at the correct anatomical region (Fig. 5A). The number of predicted possible lesion areas were high in first few iteration. Still, as the model processed more data, e.g. in iters 4 and 5 no images were predicted with more than 2 ROIs (supplementary S2C). Fig. 5C depicts the detection rate of each image across iteration, i.e. 122 images were not detected/predicted with any lesion in any of the training iters and 145 images were consistently predicted with lesion masks with models from all 5 iterations.

**Figure 5:**
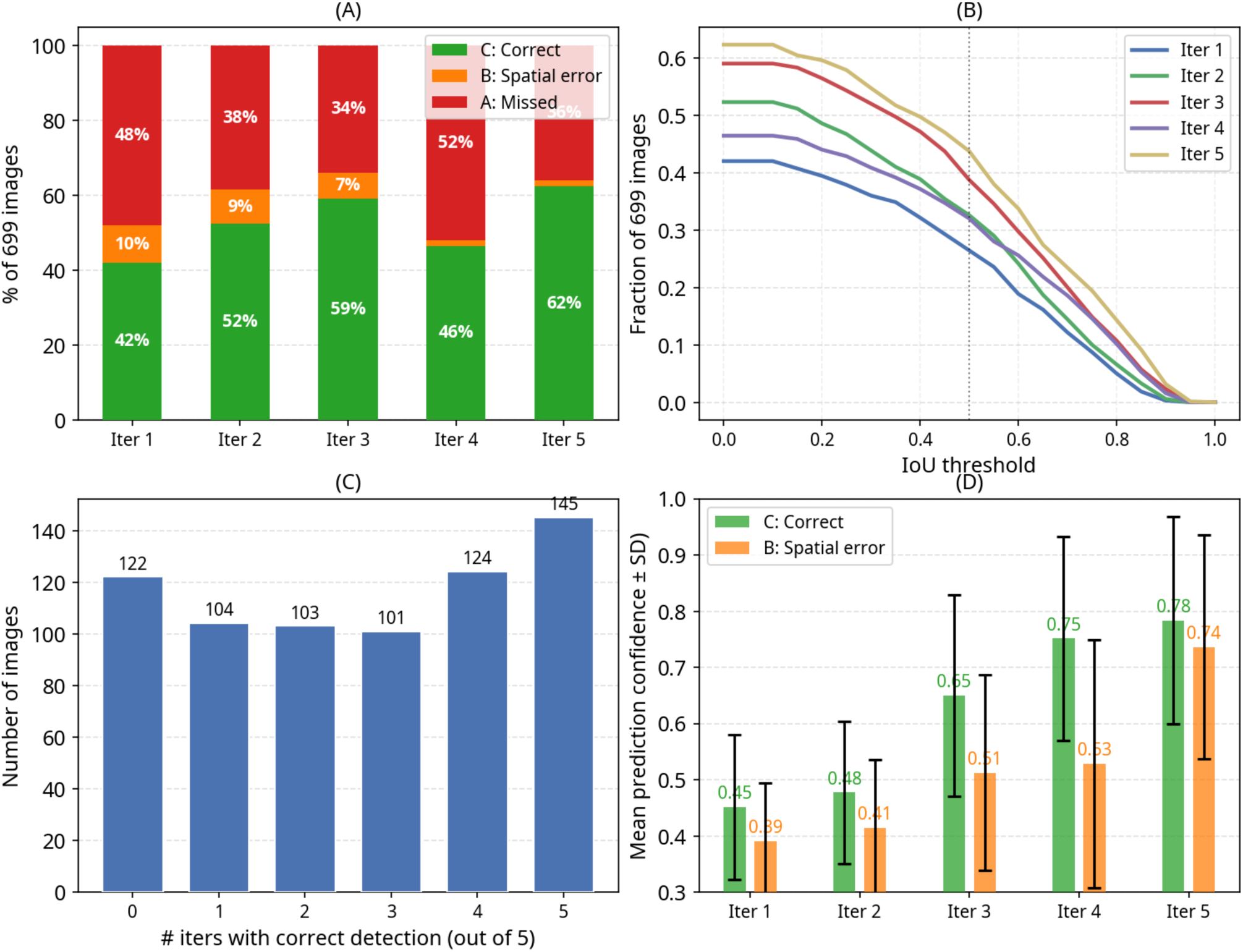
Insight into the segmentation outcomes - breakdown, segmentation quality, and confidence. *Panel* (A) shows the dramatic drop in spatial errors (Case B, orange) from 10% in iters 1 → 2 to 2% in iter 4-5, the model stops hallucinating lesions in wrong locations. (B) Fraction of Images with maxloU *>* threshold (denominator = all 699 GT-positive images). (C) per-image consistency, e.g. 145 images were always correctly detected across all 5 iters and 122 were never detected in any. (D) shows that by iter 4, spatial-error predictions have nearly as high a mean confidence as correct ones, indicating that the confidence score cannot serve as the sole judge.

**Table 3:** Detection outcome breakdown across five HITL iterations on the 699-image internal test set. All images are GT-positive. Case A = no prediction; Case B = prediction made but no GT polygon matched (IoU *<* 0.1); Case C = at least one GT polygon matched. maxIoU and maxDice are reported only for Case C images.

| Iter | Train (mask) | Case A | Case B | Case C | Mean $\text{maxIoU}$ | Mean $\text{maxDice}$ | Med. conf |
| --- | --- | --- | --- | --- | --- | --- | --- |
| 1 | 233 | 336 (48.1%) | 69 (9.9%) | 294 (42.1%) | 0.558 | 0.693 | 0.43 |
| 2 | 474 | 269 (38.5%) | 64 (9.2%) | 366 (52.4%) | 0.549 | 0.684 | 0.47 |
| 3 | 973 | 237 (33.9%) | 49 (7.0%) | 413 (59.1%) | 0.587 | 0.716 | 0.69 |
| 4 | 1907 | 363 (51.9%) | 11 (1.6%) | 325 (46.5%) | 0.599 | 0.725 | 0.80 |
| 5 | 2705 | 252 (36.1%) | 11 (1.6%) | 436 (62.4%) | 0.601 | 0.725 | 0.86 |

#### 2.3.2 Segmentation quality

Among Case C images, both mean maxIoU and mean maxDice increased across iterations (Fig. 6 & Fig. 5B). Mean maxIoU rose from 0.558 (iter 1) to 0.601 (iter 5), and mean maxDice from 0.693 to 0.725 over the same range (Fig. 6B). Notably, the quality improvement was monotonic even during iter 4, when the detection rate temporarily declined: although fewer images were correctly detected in iter 4 (Fig. 5A, 8A), the segmentation boundary precision among those that were detected was higher than in any earlier iteration (mean maxIoU 0.599, mean maxDice 0.725). The IoU threshold sweep (Fig. 5B) illustrates this clearly: at IoU *>* 0.5, iter 5 detected 43.8% of all 699 images (306/699), compared to only 26.5% (185/699) in iter 1.

**Figure 6:**
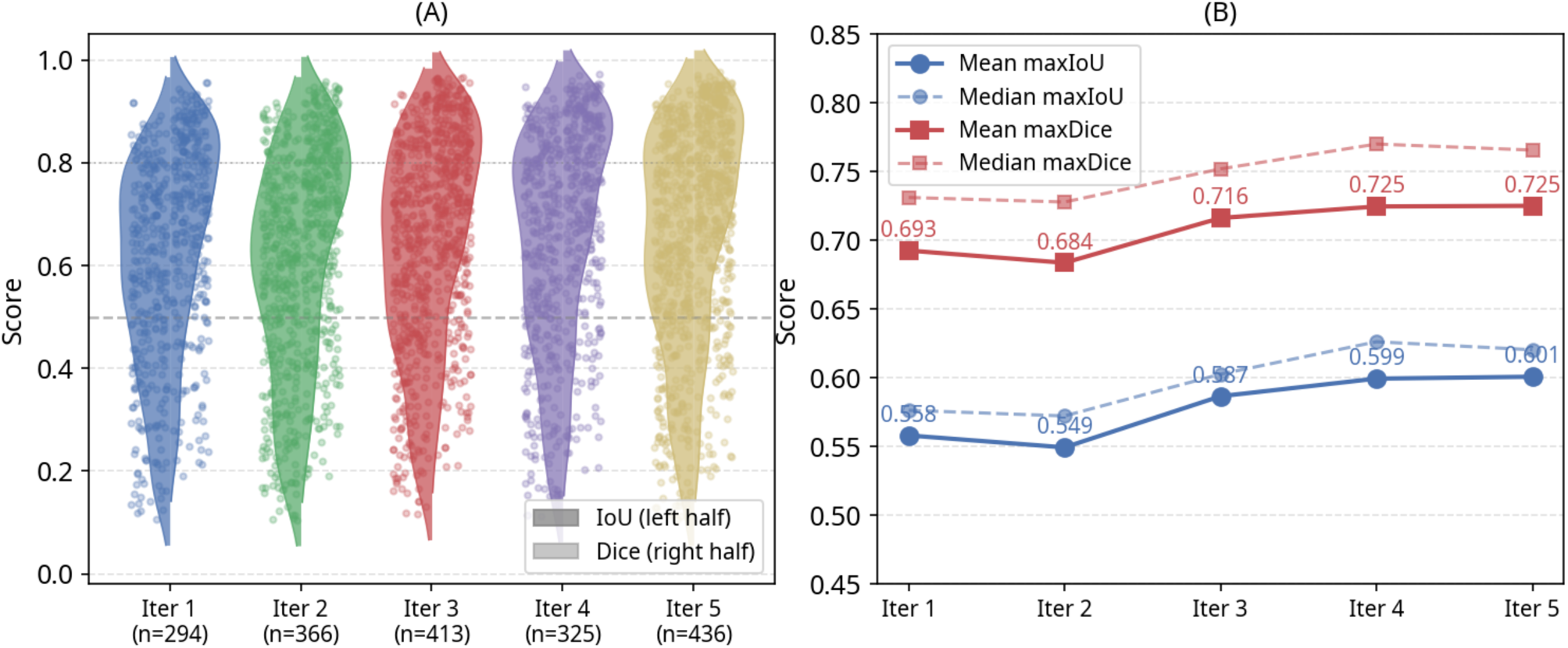
Insight into the segmentation quality across HITL training: *Panel* (A) shows the IoU and Dice score distribution across iterations among Case C (correct prediction with substantial overlap with GT). (B) mean and median maxloU / maxDice Trend (matched images only).

#### 2.3.3 Model confidence

The top-1-YOLO confidence score (the top IoU prediction mask in correctly predicted images above an IoU threshold, with one or more predicted masks) increased substantially and monotonically across iterations (Fig. 7). The distribution shifted from a median of 0.43 in iter 1 (nearly all predictions below 0.7, none above 0.8) to a median of 0.86 in iter 5. The majority of predictions were above 0.8, with a sharp KDE (Kernel Density Estimate) peak over 0.9 (Fig. 7A) for iter 5. This shift is consistent across all five prediction slots (Fig. 7B), indicating that the increase in confidence reflects a genuine model-level change rather than a selection artifact.

**Figure 7:**
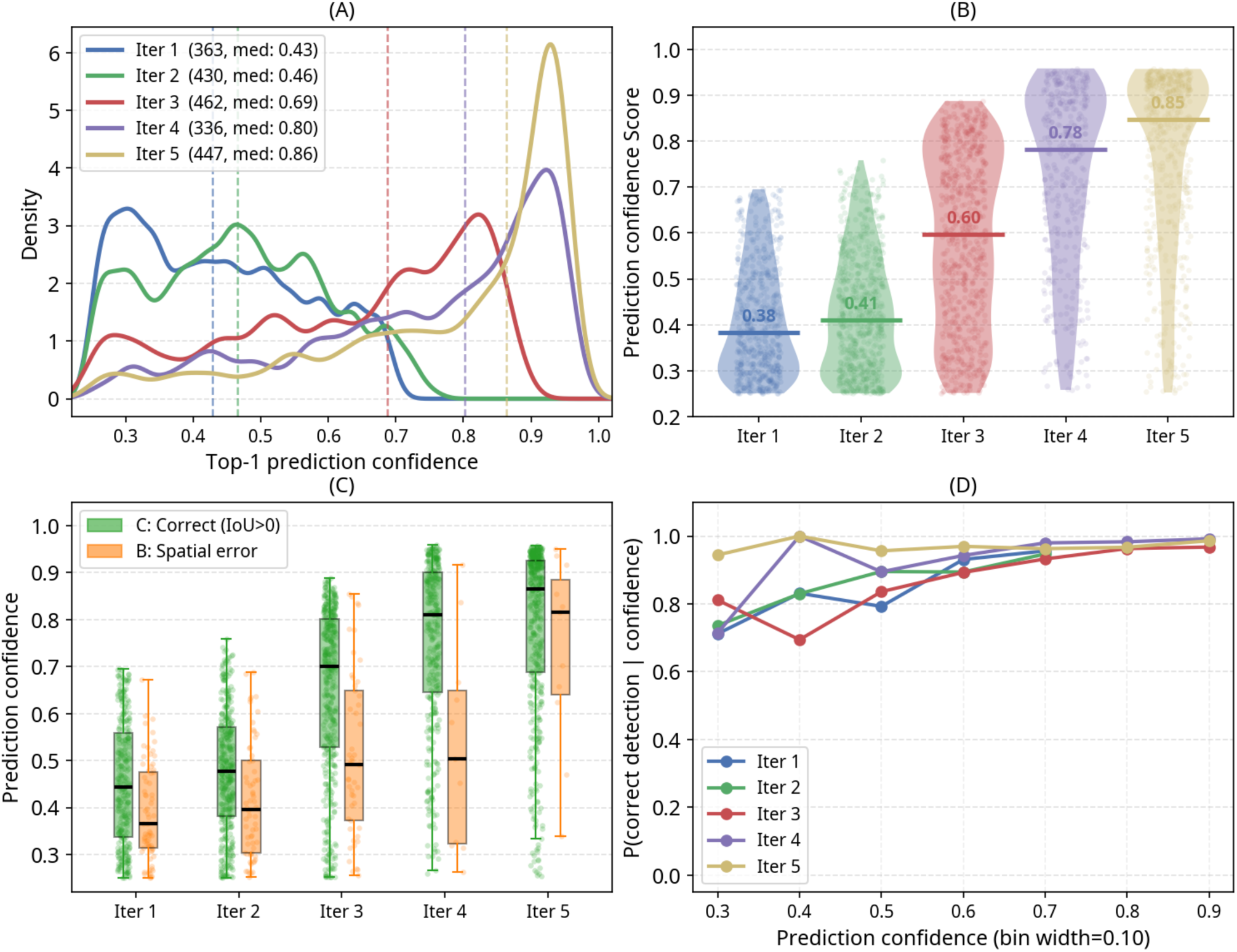
Model confidence and prediction correctness across iterative pseudo-label training (for n=699 GT-positive images): *Panel* (A) The KDE estimate of the prediction confidence (Top-1) distribution; (B) Change in distribution of confidence scores across iterations.Model prediction (classification and segmentation) vs. confidence across iterative pseudo-label training (699 GT-positive images): (C) Confidence distributions (box + jitter) stratified by prediction outcome (Case C: correct, Case B: spatial error) per iteration;(D) Empirical probability of correct detection (Case C) as a function of binned yoloConfidence1 (bin width = 0.10) per iteration.

#### 2.3.4 Confidence versus correctness

Fig. 7C shows the confidence distributions of Case C (correct) and Case B (spatial error) predictions across the five iterations. In iters 1 and 2, the median confidence of Case B predictions (≈ 0.35–0.40) was substantially lower than that of Case C predictions (≈ 0.45–0.50), indicating that the model expressed genuine uncertainty about its spatially displaced predictions in the early iterations. By iters 4 and 5, however, this separation largely disappeared: Case B predictions rose to medians approaching those of Case C (both near 0.85 in iter 5), signaling a progressive loss of confidence-based discriminability between correct and erroneous predictions. This calibration degradation is discussed further in discussion.

Fig. 7D reveals a complementary and perhaps counter-intuitive pattern: in iters 1-3, the empirical probability of a correct detection (Case C) shows meaningful variation across confidence bins, with low-confidence predictions being somewhat less reliable. By iters 4 and 5, however, the curves compress and flatten near *P* = 1.0 across virtually all confidence bins. This reflects the near-disappearance of Case B events by iter 4 (1.6% of predictions) rather than a meaningful improvement in the confidence-correctness relationship: almost every prediction made is spatially correct, regardless of its assigned confidence score, so the conditional probability P(correct *|* confidence) is uniformly high. Confidence has thus become uninformative as a discriminator, not because the model is well-calibrated, but because the few remaining spatial errors carry equally high confidence to correct predictions.

Fig. S3A, B show the scatter of yoloConfidence1 against maxIoU and maxDice, respectively, for Case C images only. The Pearson correlation between confidence and maxIoU declined monotonically from *r* = 0.46 (iter 1) to *r* = 0.17 (iter 4), with a marginal recovery to *r* = 0.22 at iter 5; a parallel decline was observed for maxDice (*r* = 0.43 → 0.16 → 0.20). Critically, this declining correlation does not indicate worsening segmentation quality - maxIoU and maxDice themselves increased across iterations. Rather, it reflects a rising floor: in later iterations, even low-confidence predictions among Case C images achieve relatively high IoU, shifting the OLS trend line upward and compressing the dynamic range of the relationship. The model increasingly achieves good boundary localization regardless of its confidence so that confidence no longer carries information about segmentation precision within the correctly detected subset.

#### 2.3.5 Per-image consistency

Of the 699 images, 122 were never correctly detected (Case C) across any of the five iterations, 145 were correctly detected in all five iterations, and 432 showed variable detection across iterations (Fig. 5C, x=0). The 145 consistently detected images (Fig. 5C, x=5) show monotonically improving maxIoU across iterations (Fig. S2B), with median maxIoU rising from 0.67 (iter 1) to 0.73 (iter 5), indicating that even on already-detected images, the model’s spatial boundary quality continued to improve. The 122 persistently undetected images represent the hard cases discussed in the next section.

#### 2.3.6 Effect of GT annotation quality

Stratified analysis (Fig. 8) reveals a consistent performance gap between Expert GT images and Pseudo-label GT images across all five iterations. Correct detection rates in iters 3-5 were approximately 5-12 percentage points higher for Pseudo-label GT images than for Expert GT images (Fig. 8A). Mean maxIoU and maxDice against Expert STAPLE GT were systematically lower than against Pseudo-label GT across all iterations; the metric distribution for case C images also depicts a similar pattern in the violin plots (Fig. 8B and C). The miss rate (Case A) was consistently higher for Expert GT images (Fig. 8D).

**Figure 8:**
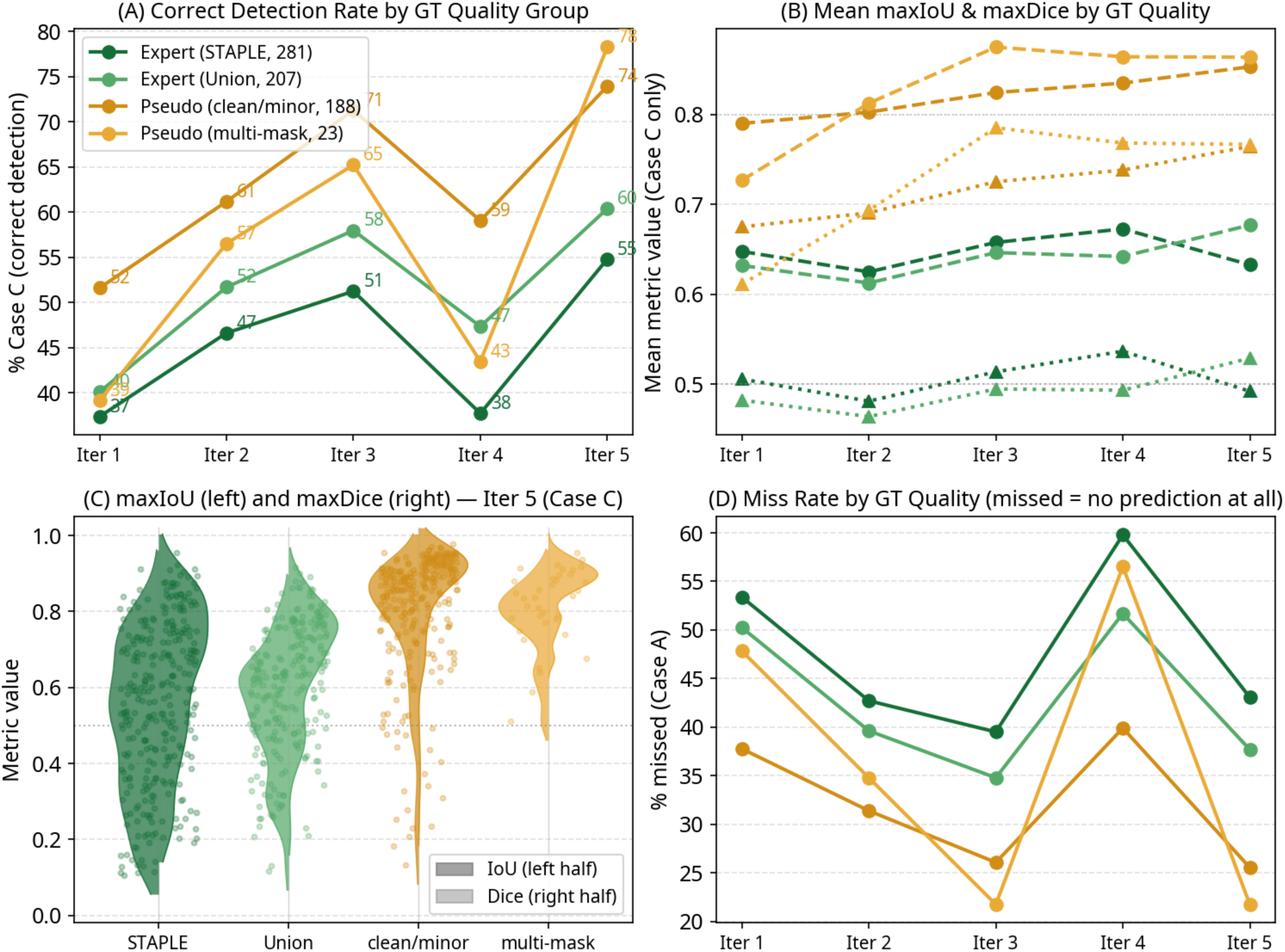
Performance of the segmentation model stratified by GT Quality (Expert vs Pseudo-Label), Expert-labels: STAPLE (n=281), Union (n=207); Pseudo-labels: clean (n=188), multi-mask (n=23): (A) Correct Detection Rate by GT Quality: Fraction of Images with maxloU *>* threshold (denominator: all 699 GT-positive images); (B) Mean maxIoU/maxDice trend across iterations among Case C (correct prediction with substantial overlap with GT; (C) maxIoU and maxDice distribution violin (Iter 5 only); (D) Miss Rate by GT Quality.

### 2.4 Reviewer Agreement Analysis

The dataset was filtered to exclude samples with known tagging errors or duplicates (leading to *N_clean_* = 787), for the RA agreement analysis 4.2.6. Pixel-wise Cohen’s Kappa was computed for all reviewer pairs. Bootstrapping (*n* = 1000 resamples) was used to generate 95% Confidence Intervals (CIs) for pairs with sufficient overlap [30, 31].

The results demonstrate a robust consensus among the primary annotators (RA3, RA2, and RA1) (Table 4). The pairwise comparisons between the three raters yield substantial agreement, with mean *κ* values ranging from 0.635 to 0.652. The tight 95% confidence intervals (e.g., RA3-RA1: [0.637, 0.669]) indicate statistical stability across the large sample size (*N >* 500 for these pairs). RA4 appears in significantly fewer samples (*N* ≤ 8 across pairs). While the RA3-RA4 and RA2-RA4 pairs show comparable *κ* values to the main group, the RA4-RA1 pairing shows a notably lower agreement (*κ* = 0.431) with a wide confidence interval, mostly due to a statistically insignificant number of samples.

**Table 4:** Pairwise Inter-Rater Reliability Statistics (Cohen’s Kappa and Observed Agreement)

| Rater Pair | Count ($N$) | Mean $\kappa$ [95% CI] | Mean Agree (%) |
| --- | --- | --- | --- |
| RA3-RA2 | 558 | 0.635 [0.617, 0.650] | 96.0% |
| RA3-RA1 | 622 | 0.652 [0.637, 0.669] | 97.0% |
| RA2-RA1 | 527 | 0.648 [0.629, 0.664] | 96.1% |
| RA3-RA4 | 8 | 0.562 [0.332, 0.721] | 98.3% |
| RA2-RA4 | 2 | 0.661 $N/A$ | 98.0% |
| RA4-RA1 | 6 | 0.431 [0.179, 0.651] | 98.2% |

## 3 Discussion

The development of deployable, smartphone-based AI screening tools for oral cancer in low-resource settings is fundamentally constrained by the lack of high-quality, pixel-level annotated data. In this study, we addressed this challenge by designing, implementing, and empirically validating a DL-driven HITL The experiment on the presence of noisy label in training yields two complementary conclusions. First, a moderate fraction of unreviewed mid-pipeline pseudo-labels (∼ 10%) does not catastrophically degrade aggregate segmentation performance, which is a reassuring property for HITL annotation frameworks in settings where expert review capacity is limited [32]. Second, and more consequentially, even this modest noise fraction introduced measurable hidden costs: a clinically unfavorable shift toward lower recall, and a three-to-four-fold increase in training instability as measured by cross-fold Ep_best_ variance. Together, these findings retrospectively validate two design decisions of the annotation pipeline (Section 4.2): (i) the investment in medical expert review of all pseudo-labels before inclusion in final training, and (ii) the use of iterative HITL cycles to progressively improve label quality rather than accepting all pseudo-labels from any single iteration. The fact that these 119 files were ultimately reviewed by three clinical reviewers and corrected where necessary confirms that the HITL protocol already addressed this source of noise; the present experiment quantifies what would have been forfeited had that review step been skipped. The convergence instability finding has a further implication for the iterative training described in Section 2.3: the non-monotonic detection rate trajectory observed on the 699-image internal test set, specifically the recall dip at iteration 4 is consistent with a scaled version of the same mechanism, where increasingly ambiguous pseudo-labels from the HITL long-tail introduced gradient noise analogous to, but more diffuse than, the controlled 119-label perturbation studied here.

The aggregate robustness of both box and mask mAP50-95 across models and datasets (with or without noisy 119 labels, see Table 2) is noteworthy. Because the 119 pseudo-labels originated from a model that had already undergone several training iterations, the annotation errors were likely subtle – boundary imprecision, minor localization shifts, or occasional spurious stray regions, rather than gross mislabeling. Such mid-pipeline pseudo-label noise is known to be less disruptive than early-iteration or randomly corrupted labels because the erroneous masks still capture the correct anatomical region at a coarse level[32, 33]. The finding also aligns with the broader literature showing that deep neural networks (DNNs) trained from scratch can tolerate noise rates below approximately 10–15% without catastrophic performance degradation on aggregate metrics, particularly when the noise is instance-level (spatial boundary errors) rather than category-level[33].

The precision-up, recall-down signature is consistent with a conservative prediction bias induced by a noisy training signal: the model learns to suppress borderline detections in ambiguous boundary regions that co-occur with uncertain pseudo-labels, thereby avoiding false positives at the cost of sensitivity [32, 33]. In the context of oral lesion screening – where failing to detect a true lesion (false negative) is clinically more detrimental than a spurious detection, even a 1.64pp reduction in mask recall is a substantive concern and independently motivates the removal of unreviewed pseudo-labels from training, regardless of the mAP-level outcome.

The most pronounced and practically significant difference between the noisy and clean configurations was not in the final metric values but in the *stability of convergence*. The wide range of Ep_best_ in the imperfect condition - spanning epoch 185 to 847 for box detection across five folds, reflects the DNN memorization phenomenon: networks initially learn clean, consistent patterns and later begin fitting noisy samples; but the epoch at which this transition begins, depends on which noisy samples appear in each training partition [33]. For instance segmentation specifically, pixel-level boundary noise in even a small fraction of labels can corrupt the fine-grained spatial gradients that govern the mask head, producing a noisier loss surface and unpredictable convergence trajectories[32]. High Ep_best_ variance also has direct practical consequences: it renders early stopping less reliable as a regularisation device and makes computational cost unpredictable across runs.

Interestingly, the elevated cross-fold variance in Ep_best_ can itself serve as a diagnostic signal for label contamination. Recent work by Chen et. al. has formalized this intuition, demonstrating that samples which consistently contribute to inferior cross-validation performance can be identified as likely noisy labels through repeated cross-validation without requiring a separate clean reference set[34].

Next, our empirical analysis of the pipeline’s learning dynamics yielded several critical insights for medical AI annotation workflows. The core observation from this analysis is that iterative pseudo-labeling – conducted primarily by a non-clinical AI-ML scientist selecting plausible predictions - produced measurable and sustained improvements in both detection rate and segmentation quality over five iterations. This is consistent with the central premise of semi-supervised self-training: that a model trained on even imperfect pseudo-labels can discover better internal representations than one confined to a small labelled set, provided the pseudo-labels carry some structural information about the target[35–37]. Specifically, iteratively generating pseudo-labels and training the model can progressively leverage information from unlabelled data to improve segmentation performance, even when those labels are imperfect.

The confidence trajectory can particularly be informative as a proxy for the model’s internal state. The core principle of self-training involves iteratively assigning pseudo-labels to unlabeled samples with confidence scores above a certain threshold, enriching the labeled dataset and retraining the classifier. In our pipeline, no explicit confidence threshold was applied during pseudo-label selection - the triage step was performed by a human reviewer who examined visual plausibility. However, the confidence scores produced by the trained models serve as a retrospective indicator of the quality of the training signal received. The shift from a flat, low-confidence distribution in iter 1 (median 0.43) to a high-peaked distribution in iter 5 (median 0.86) mirrors what is theoretically expected from iterative self-training: as each model generates higher-quality pseudo-labels for the next training round, the successor model encounters less conflicting gradient signal and converges to more decisive feature boundaries [38].

The saturation and partial reversal in detection rate during iter 4, followed by recovery in iter 5, is consistent with a well-described phenomenon in iterative pseudo-labeling: as the training set expands to include harder, more ambiguous cases, which constitute the long tail of the dataset - the model temporarily loses recall on easier images because it must reconcile contradictory spatial signals from different subsets [36, 37]. The observation that segmentation quality (maxIoU and maxDice) continued to improve monotonically even during this recall dip suggests that the model had internalized better boundary representations; it simply became more conservative about when to predict them.

The most noteworthy finding is that substantial improvements in segmentation quality were achieved across all five iterations without any expert clinical input beyond the initial 233-image seed set. The correct detection rate increased from 42.1% (iter 1) to 62.4% (iter 5), and mean maxDice improved from 0.693 to 0.725 among detected images, purely through AI-ML-guided triage of pseudo-labels. This aligns with the broader finding in the pseudo-labeling literature that *data reinforcement helps, even with error-prone pseudo-labels*, and that Convolutional Neural Network (CNN)s possess a natural regularization capacity with respect to labeling errors, provided the noise rate remains below a critical threshold [37]. In our pipeline, the human reviewer’s role – rejecting visually implausible predictions (reflections, non-oral regions, clothes), effectively served as a coarse quality gate without requiring clinical expertise, demonstrating the viability of a non-expert’s (NMA) contribution in the HITL loop in the early and middle phases of dataset construction.

From a practical standpoint, this result is encouraging for resource-limited settings where clinical expert capacity is scarce [39]. It suggests that a mixed strategy - expert annotation for a small high-quality seed set combined with non-expert-guided pseudo-label expansion, can yield progressively improving models, with clinical review concentrated at the final quality assurance stage [40] rather than distributed across all iterations.

Despite the overall positive trend, two distinct performance gaps warrant careful interpretation. First, 122 images (17.4%) remained in Case A across all five iterations; the model never generated a prediction at all for them. Inspection of the gtCount distribution suggests these are not disproportionately multi-lesion images; more likely they represent visually atypical presentations (e.g., very early-stage or low-contrast lesions, heavily occluded views, or images acquired under poor illumination) for which the model has not yet encountered sufficient training analogues. This is consistent with the theoretical prediction of the *confirmation bias* in pseudo-labeling: once a model consistently fails to predict a lesion, that image is never selected as a pseudo-label, so the model never receives any corrective gradient signal for that type of appearance [35, 41, 42]. This situation is reinforced when incorrect predictions are used as labels for unlabelled samples, as it is the case in pseudo-labeling. The 122 images that were persistently missed, represent a fundamental blind spot of the iterative self-training approach in the absence of active learning, contrastive learning, or targeted annotation strategies as in text-data domains [43].

Secondly, the GT quality stratification shows that the model performed systematically better on pseudo-label GT images than on expert-annotated (STAPLE and Union) GT images across all iterations. Two mechanisms contribute to this. The first is:

- *distribution alignment*: the training pseudo-labels and the test pseudo-label GT share the same annotation style (single clean polygon following the lesion contour) and even the same generating process (morphological opening + contour extraction from a YOLO prediction), so the model’s output closely matches the shape and spatial extent of the test GT for these images. This is a form of in-distribution advantage analogous to the domain shift between training and unseen test sets.
- The second mechanism is genuine *GT* difficulty: STAPLE-consensus masks, derived from three independent expert annotations, are spatially smoother and often broader than the model’s polygon output, as the consensus algorithm probabilistically enlarges the mask to encompass the region agreed upon by multiple annotators. The Union-based masks for the two-reviewer subset are similarly expanded. A lower maxIoU against expert GT does not necessarily indicate a worse predictions; it may reflect the tighter coverage expected from a model’s output compared to a human-drawn consensus boundary.

This is the direct analogue of the systematic bias observed in lesion segmentation benchmarks, where models trained and evaluated under different annotation protocols exhibit metric discrepancies that do not reflect differences in clinical accuracy [33]. Final evaluation should therefore separately report metrics against stratified sets - expert GT (reflecting clinical ground truth quality) and against pseudo-label GT (reflecting self-consistency of the pipeline), even when both are reviewed by domain experts (as in our case).

In regards to the model prediction confidence, the declining Pearson correlation between confidence and maxIoU across iterations (*r* = 0.46 → 0.22) deserves careful interpretation. It does *not* indicate that the model’s segmentation quality deteriorated; maxIoU itself increased. Rather, it reflects a progressive decoupling of confidence from segmentation precision: later models become uniformly high-confidence regardless of whether they achieve high or low IoU on a given image. This is a recognized form of *overconfidence* that arises in iterative self-training when the model receives a sustained stream of pseudo-labels that it generated itself - a reinforcing loop in which the model is never confronted with confident predictions that were subsequently labeled incorrect by an external authority [41]. The manifestation in Fig. 5D is that by iter 4-5, Case B (spatially erroneous) predictions have nearly the same mean confidence as Case C (correct) predictions, indicating that confidence alone can no longer reliably distinguish correct from incorrect predictions. In a clinical deployment context, where confidence scores might be used to triage images for human review, this calibration degradation would be consequential. Temperature scaling or Platt calibration on a small calibration set could be the a mitigation strategy and is recommended before any downstream clinical use of the model[44].

Most of the images in our set had single annotation mask, but a small set of images had multiple disconnected masks. The GT lesion count (gtCount) analysis (Fig. S2A) shows a consistent decline in the correct detection rate as gtCount increases, present across all five iterations. For images with a single annotated lesion (gtCount=1, n=605), correct detection rates ranged from approximately 47% (iter 1) to 65% (iter 5). For images with two or more annotated lesions (gtCount ≥ 2, n=94), rates were markedly lower across all iterations. Two factors compound in multi-lesion images: the greedy matching framework may pair a prediction to a secondary lesion while the primary lesion goes undetected, and the model –trained predominantly on single-lesion pseudo-labels (by distribution, and also in some cases, because the morphological opening step tended to retain only the largest component) – may not have internalized the spatial understanding of multi-lesion presentations. Such multi-instance examples are known to require instance segmentation architectures to handle correctly [45]. The YOLO model in principle supports multi-instance prediction, but its training distribution here was biased toward single-instance outputs, which explains this observation.

Quantifying the agreement between reviewers and comparison with the AI-performance yield valuable insights. According to the Landis and Koch interpretation scale [46], values between 0.61 and 0.80 represent “Substantial Agreement,” validating the consistency of the segmentation protocols for the majority of the dataset, particularly where the DL model failed to generate acceptable annotations. A notable observation is the discrepancy between the extremely high Raw Agreement (*>* 96%) and the moderate Kappa values (≈ 0.65). This phenomenon, known as the *prevalence paradox*, occurs in medical imaging when the region of interest (lesion) is small relative to the background[30, 47, 48]. In such cases, raters easily agree on the large background area (*D*), inflating *p_o_*. Cohen’s Kappa successfully corrects for this by penalizing the score based on the probability of chance agreement, providing a more rigorous assessment of the actual lesion segmentation overlap than accuracy alone[46, 49]. The statistical divergence (low R4-R1 agreement) is likely statistically insignificant, given the low sample count (*N* = 6), but such scenarios warrant manual review of those specific instances to ensure consistent annotation standards.

The systematic gap between model performance against Expert GT (STAPLE/Union) versus Pseudo-label GT, is interpretable in light of the inter-rater reliability analysis. The pairwise Cohen’s *κ* values of 0.635–0.652 between the primary reviewers indicate that even among experienced clinicians, substantial pixel-level disagreement persists on these images. Yang et al. similarly observed, for a comparable binary segmentation task (chest radiograph abnormalities), that expert pairwise IoU against STAPLE consensus was 0.54 on average – closely matching our model’s mean maxIoU of 0.55–0.60 against Expert GT [50]. This suggests that the model’s Expert-GT performance is approaching the human inter-annotator ceiling for this class of visually ambiguous lesion boundary tasks, rather than indicating a systematic model failure. The pseudo-label GT advantage is an in-distribution artifact, not a reflection of better absolute quality.

The substantial agreement (*κ* = 0.635–0.652) observed between the three primary reviewers defines an interpretable upper bound for model-vs-GT performance metrics. A DL model evaluated against a single reviewer or consensus GT cannot be expected to exceed the agreement level of a second human reviewer against the same GT [50, 51]. Critically, the mean IoU between expert annotators and the STAPLE consensus (≈ 0.54 in analogous tasks [50]) is numerically similar to the mean maxIoU achieved by the later HITL models against Expert GT on the 699-image test set (0.599–0.601; Table 3). This convergence implies that the model boundary predictions — at the final training iteration are operating within the inter-human agreement envelope, lending validity to the annotation pipeline as a whole [51]. Conversely, the persistently undetected images (122 of 699, never Case C across five iterations) likely represent the hardest subset of visually ambiguous cases where even human reviewers disagree on boundary placement, as evidenced by the wide confidence intervals in the R4 pairs (*κ* = 0.431, [0.179, 0.651], *N* = 6) that suggest genuine boundary uncertainty at the level of individual images. Future work could focus annotation effort on exactly this hard cohort, using targeted active learning [43, 52] to break the confirmation bias cycle.

Overall, we present a HITL workflow for OPMD lesion segmentation, with several critical insights. First, we demonstrated that iterative self-training significantly improves both the spatial localization and boundary precision of oral lesions, even when guided by non-clinical algorithmic triage in the intermediate stages. However, our controlled experiments on label noise revealed that the inclusion of even a small fraction (∼10%) of unreviewed pseudo-labels introduces substantial convergence instability and induces a conservative prediction bias. Because this bias clinically manifests as a reduction in model recall - an unacceptable trade-off in cancer screening, our findings underscore the absolute necessity of integrating a final, expert-driven quality assurance step within the HITL loop. Our analysis revealed that the segmentation model’s performance against the multi-expert consensus ground truth effectively mirrors the inter-rater reliability ceiling (*κ* ≈ 0.65). This convergence indicates that the model’s boundary predictions fall within the expected inter-human agreement envelope, robustly validating the overall efficacy of the HITL annotation pipeline.

Despite these successes, our study highlights important limitations that define future research directions. We observed a progressive decoupling of model confidence from segmentation accuracy in later HITL iterations. Additionally, a persistent subset of visually atypical and multi-lesion images evaded detection entirely, demonstrating that passive pseudo-labeling is insufficient for the most challenging edge cases - necessitating the incorporation of targeted active learning strategies in future iterations of this pipeline.

Ultimately, this work provides a systematically validated blueprint for curating large-scale, domain-specific medical imaging datasets. By successfully balancing the scalability of deep learning with the rigorous oversight of clinical experts, this HITL framework bridges the gap between raw clinical images and deployable AI, laying the methodological groundwork for point-of-care oral cancer screening in the populations that need it most.

## 4 Methodology

### 4.1 Image source and annotators

The images were collected between 2010 and 2022 (over a decade) through public health efforts by the Biocon Foundation (https://bioconfoundation.org). The images were taken from habit-positive (history of smoking and/or chewing tobacco or its derivatives) individuals with self-reported oral lesions, who were at least 18 years or older. The image capture devices were low- to mid-range smartphones with at least a 5-megapixel camera, such as the Motorola Moto G5 and iQOO. The images after anonymization, were of different sizes or resolution with the minimum being 1024 × 709 and maximum 4608 × 3456. Three different service providers provided technical support with a signed MoU with the Biocon Foundation. All regulatory approvals and compliance requirements were adhered to in these oral cancer screening initiatives. The data for this study were made available through a Data Sharing and Collaboration Agreement between the Biocon Foundation and IISc under agreement number IN-KA89771284129871W.

At the triage stage, reviewer-cum-annotator (RA) with 10 (RA1) and 14 (RA2) years of experience in Oral Medicine and Radiology were engaged independently for labeling and drawing freeform masks (pixel-level annotation) of lesions in the OPMD images. They reviewed all the images with the lesion mask predicted by our DL model, and manually annotated them when deemed necessary as per their assessment. A third (RA3) and a fourth (RA4), with 6 and 25 years of experience, respectively, reviewed and annotated the images, whose annotations from the first two were widely discordant, and at times even for class labels (suspicious and non-suspicious) that did not match.

### 4.2 AI-ML iterative pipeline with HITL

#### 4.2.1 Stage 1: Iterative pseudo-labeling prior to expert review

We utilized a DL algorithm for oral cancer lesion annotation, incorporating the HITL approach to integrate expert knowledge and guidance. Briefly, our starting dataset consists of 4887 mobile phone images (Set A), labeled by telespecialists as potential OPMDs having suspicious lesions. RA1 and RA2 manually marked the original ROIs having suspicious lesions for 233 images. We initiated the process with the pairs of images of the oral cavity and corresponding annotation masks (labels), to fine-tune a pre-trained Convolutional Neural Network (CNN)-based model (yolo11x-seg). This model had been pre-trained on the COCO dataset providing a good overall understanding of human facial features [28]. This fine-tuned model was subsequently used to predict annotations on Set A, and it predicted masks for 2020 images (Fig. 2). We (non-medical annotator (NMA)) reviewed all images and identified only 241 masks (pseudo-labels) suitable for next model training. In the next iteration, a total of 474 (233+241) images were used to train the COCO-pre-trained model from scratch and this model could predict masks for 2411 images, from which 499 were selected for the next training set along with the previously culled 474 images. Our iterative procedure went on until we had a majority of the images annotated. Until this point, the model had human input only for lesion localization and error mitigation. For example, at the initial stages, the model frequently predicted lesions at locations with reflection, regardless of whether a lesion was present. Also, the model marked irrelevant areas, including extra-oral regions, appendages aiding oral examination, clothes, etc. All those images were kept outside of the training, and only those images were selected where a lesion could be selected from straightforward examination. At the 5th iteration, the model showed signs of learning saturation with no improvement in detection rate (Table 1), and 794 images with no mask prediction remained in set A. At this stage, all images of set A and pixel annotations (also known as pseudo-labels) generated by the DL model were sent to the RAs for review, and manual annotation. Scrutiny revealed that 1198 images had distinct characteristics, were captured using a dual-modality (autofluorescence) imaging device, and were not being segmented by the model. These images were kept aside. And of the remaining 3689 images, 535 were flagged as non-suspicious, 128 were excluded (blur or other artifacts rendering them as non-diagnosable) by at least two RAs, hence eliminated from the data set leaving 794 images for manual annotation. This yielded a total of 3026 images for HITL annotation.

#### 4.2.2 Stage 2: Expert review, annotation, curation

The model generated annotation masks on a subset of images, which were then sent to RA1 and RA2 for individual independent review. Each RA provided one of the four responses for each image and manually annotated as needed:

- *Agreement (ok):* The RA agrees with the model-predicted Region of Interest (ROI). That is, the DL-model predicted pseudo-label (annotation mask) is properly localizing and denoting the lesion boundary, and no need for modifying the mask.
- *Disagreement (notOk)*: RA deemed the pseudo-label as incorrect or of insufficient quality - in this case the reviewer manually re-drew the ROI (using LabelMe software); https://labelme.io).
- *Non-diagnosable (exclude)*: RAs cannot diagnose the image due to poor quality; the image is removed from the annotation/curation pool.
- *Non-suspicious (ns)*: RA suggested the image contains no suspicious lesion(s); the image is moved to the non-suspicious pool and excluded from further training.

We maintained at least two RAs for each image. In cases of discordance or a bias was suspected, the image was sent to a third RA (RA3), or even a fourth (RA4), who possessed the most experience in Oral Medicine and Radiology. These corrected annotations were utilized for post-processing and training the updated segmentation model.

#### 4.2.3 Stage 3: Collating the whole set and post-processing

Following review, we merged the ‘ok’ images with the original manually annotated images to iterate the DL fine-tuning from scratch and predict again on Set A. By repeating this process multiple times, we obtained a higher quantity and quality of annotated images in each iteration, until the majority of suspicious images were annotated properly as per expected quality.

##### 4.2.3.1 Post-processing of YOLO segmentation predictions via morphological opening

A custom batch post-processing pipeline was applied to the YOLO segmentation predictions for all 2,232 images (see Fig. 2 for analysis stages). For each image, the predicted polygon annotation was read from the corresponding YOLO-format TXT file, where coordinates are stored as normalized (x, y) pairs relative to image dimensions. Each polygon was rasterized onto a binary mask of the same resolution as the source image using PIL’s (https://pillowpython.com) polygon drawing routine, with all polygon instances within a given file composited onto a single mask.

Morphological opening was then applied to each binary mask using SciPy’s ‘ndimage‘ functions. The operation consisted of binary erosion followed by binary dilation, each with a 3×3 structuring element (full connectivity) and a single iteration. Morphological opening removes small spurious foreground regions and disconnected noise pixels that do not survive erosion, while largely preserving the geometry of larger structures. This step was intended to refine the boundaries of predicted segmentation regions and eliminate isolated prediction artifacts introduced by the YOLO model.

Following opening, the connected components of the resulting mask were labeled and their individual pixel areas were computed and ranked in descending order (up to 20 components recorded per image). External contours were then extracted from the opened mask using OpenCV and sorted by area. The refined annotations were saved in YOLO-format polygon TXT files with one contour per line. Summary statistics including the number of connected components, total foreground pixel area, and per-component pixel areas were recorded for each image and exported to a CSV (comma-separated values) file for downstream analysis.

##### 4.2.3.2 Manual annotation

A little more than 25% (794, Fig. 2) of the whole image set had to be manually redrawn because the DL model did not predict any lesion mask or the predictions were not accepted by the reviewers as sufficient quality. This set was first annotated by two RAs independently; we collated a total of 325 images, having two polygon masks. Here, we combined the two available masks for each image using union to generate the final image mask. For a total of 469 images, we additionally received input from RA3 and RA4, resulting in three polygon masks for these tough-to-auto-segment images. Here, each image having three manually annotated masks, we could apply the STAPLE algorithm to obtain the final lesion mask, as the algorithm requires at least three masks per image.

##### 4.2.3.3 Geometric simplification via Ramer-Douglas-Peucker (RDP)

The polygons denoting the masks were simplified using the Ramer–Douglas–Peucker (RDP) algorithm with a tolerance of *ɛ* = 1.0 pixel. The RDP algorithm reduces a polyline (polygon contour of the lesion mask) by iteratively identifying the point with the greatest perpendicular deviation from the line segment joining the current endpoints. Points whose deviation falls below *ɛ* were discarded as redundant, while those exceeding *ɛ* were retained as structurally significant. Setting *ɛ* = 1.0 pixel was motivated by two considerations. First, contours extracted from binary masks exhibit a characteristic “staircase” artifact, in which boundaries follow the rectilinear grid of pixel edges rather than smooth anatomical boundaries. Second, at the acquisition resolution of 4160 × 3120 pixels, a single-pixel displacement corresponds to approximately 0.02% of the image width, a scale well below any clinically meaningful morphological feature.

#### 4.2.4 Effect of noisy label (annotation quality) on training

To quantify the sensitivity of the yolo11n-seg model to annotation quality within a HITL pipeline, we designed a controlled 5-fold cross-validation experiment comparing two training configurations that differed in a single respect: the inclusion or exclusion of 119 image-mask pairs whose labels were pseudo-labels generated at an intermediate HITL iteration and selected by the NMA, but not yet reviewed or corrected by any RAs at that stage. We denote these two configurations as *imperfect* (training set size ∼1,265 per fold) and *clean* (∼1,147 per fold), with the noisy subset constituting approximately 10% of the clean training set. It is important to note that these 119 files were subsequently reviewed and corrected, where necessary, by RA1, RA2, and RA3 independently, and were included in the final annotation pipeline. This experiment isolates the retrospective value of expert intervention on model convergence and performance. All other training hyperparameters, model initialization, and cross-validation splits were held constant. The validation sets differ across folds by design (each fold uses a distinct held-out partition), but are identical between the two experiments for any given fold, ensuring that performance differences reflect training-set composition alone.

#### 4.2.5 Performance of the HITL iterative pseudo-label pipeline on an independent ground truth test set

To quantify how segmentation performance evolved across the five iterative HITL training cycles, each trained yolo11x-seg model was applied to a fixed hold-out set of 699 images that had not been seen by any of the five models during training. Every image in this set has an associated ground-truth polygon mask, derived through one of the post-processing or expert-annotation routes described in Fig. 2, Sec 4.2.3.2 and 2.1: 188 images carry pseudo-labels accepted without modification, 23 carry pseudo-labels whose component selection was manually adjusted, 281 carry STAPLE-consensus expert annotations from three reviewers, and 207 carry union-consensus annotations from two reviewers. Critically, all 699 images were withheld from training at all five iterations, providing a genuinely independent test set for each successive model.

##### 4.2.5.1 Evaluation and prediction–ground truth (GT) matching

For each image, the model produced between zero and five polygon instances (stored as normalized YOLO-format coordinates). GT polygons were loaded from corresponding TXT files and denormalized to pixel coordinates. Because oral lesion images can contain *multiple* annotated regions per image, a naive one-to-one comparison of the first prediction to the first GT polygon is not valid. We therefore adopted a *greedy maximum-overlap matching* strategy, consistent with standard COCO-style evaluation[29]: an all-pairs intersection over Union (IoU) matrix was constructed between every predicted polygon and every GT polygon for that image; the pair with the highest IoU was locked and removed from subsequent consideration; and the process was repeated until no remaining pair exceeded IoU *<* 0.1. For each matched pair, both IoU and Dice were recorded. The maxIoU and maxDice values reported for each image correspond to the single best-matched pair.

During analysis, three mutually exclusive outcome categories were defined for each image:

- Case A (*missed*): the model produced no prediction at all.
- Case B (*spatial error*): the model predicted at least one polygon, but no GT polygon was matched above the IoU threshold (the prediction was spatially displaced from all annotated lesion regions).
- Case C (*correct detection*): at least one predicted polygon overlapped a GT polygon, enabling a valid segmentation quality measurement.

Because all 699 images carry ground-truth lesion masks, the detection rate is equivalent to sensitivity, and specificity cannot be computed from this set alone. Plots were generated stratifying by GT annotation quality (Expert: STAPLE and Union; Pseudo: clean/minor and multi-mask) to assess whether model performance differed depending on annotation provenance (See Section 2.3.6).

#### 4.2.6 RA agreement analysis: Cohen’s Kappa in segmentation

Standard metrics like Intersection over Union (IoU) or Dice coefficients primarily evaluate the Intersection (the lesion). However, Cohen’s Kappa differs because it also accounts for the Background (non-lesion pixels) and corrects for “Agreement by Chance” [30, 31]. The statistic is calculated as:

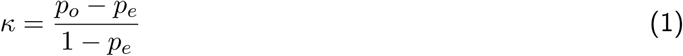

where:

- *p_o_* (Observed Agreement): The proportion of pixels where raters agree (either both classify as “Lesion” or both as “Background”).
- *p_e_* (Expected Agreement): The proportion of agreement expected by random chance, based on individual rater tendencies to mark pixels as “Lesion.”

The components are defined as:

1. A (TP): Pixels inside *both* polylines (Intersection).
2. B & C (FP/FN): Disagreement areas (where one polyline is wider or shifted).
3. D (TN): The empty space outside the polylines (Background).

Pixel-wise Application

For an image of resolution 1000 × 1000 (1,000,000 pixels), we compare two raters (e.g., R3 vs. R2) by flattening the segmentation masks into 1D arrays and constructing a pixel-wise Confusion Matrix (Table S1). The Calculation:

1. Observed Agreement (*p_o_*): 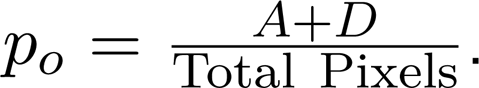 Note that since *D* (background) is typically large, *p_o_* is often very high (e.g., *>* 0.95).
2. Chance Agreement (*p_e_*): We calculate the probability of R3 saying “Yes” (*P_R_*_3_) and R2 saying “Yes” (*P_R_*_2_).

- *P_AgreeY es_* = *P_R_*_3_ × *P_R_*_2_
- *P_AgreeNo_* = (1 → *P_R_*_3_) × (1 → *P_R_*_2_)
- *p_e_* = *P_AgreeY es_* + *P_AgreeN o_*
3. *κ* Statistic: We subtract the high chance baseline (*p_e_*) from the observed agreement (*p_o_*) to normalize the score.

We used the following simple steps to obtain the 95% confidence interval (CI) via bootstrapping.

1. For any two pairs we repeatedly pick the Kappa scores with replacement (one score may appear multiple times, and one score may not appear at all) until we have the total number of available scores. Then the average score is calculated.
2. This procedure is done 1000 times yielding 1000 different averages.
3. For confidence interval (CI) calculation, the 1000 averages are sorted, and the top and bottom 2.5% cut-off values are obtained. The middle range is given as the 95% along with the average without bootstrapping.
4. Cohen’s Kappa distribution can be skewed but bootstrapping doesn’t require normal distribution assumption, so it is applicable in this case.

## Data Availability

The data for this study were made available through a Data Sharing and Collaboration Agreement between the Biocon Foundation and IISc under agreement number IN-KA89771284129871W. The data cannot be shared without prior MoU with another third party.

## Acknowledgements

We thank Department of Science and Technology, New Delhi, the Government of Karnataka, and Prof. Rajesh Sundaresan for their support. SM acknowledges the useful discussion with Pushp Lochan and Shankararama Sharma.

## Author Contributions

SM: Conceptualization, Methodology, Software, Validation, Formal Analysis, Investigation, Visualization, Data Curation, Writing – Original Draft. PM, KG, HT, PB: Data Curation, Investigation. AS: Resources. DP: Conceptualization, Writing – Review & Editing, Supervision, Project Administration, Funding Acquisition.

