## supplementary for "Human In the Loop Challenges for Quality Annotation of Pre-Cancer Lesions in Clinical Oral Images"

Table S1: Pixel-wise Confusion Matrix for Segmentation

| | **R2 says Lesion (1)** | **R2 says Background (0)** |
|---|---|---|
| **R3 says Lesion (1)** | **A** (True Positive) | **B** (R3 only) |
| **R3 says Background (0)** | **C** (R2 only) | **D** (True Negative) |

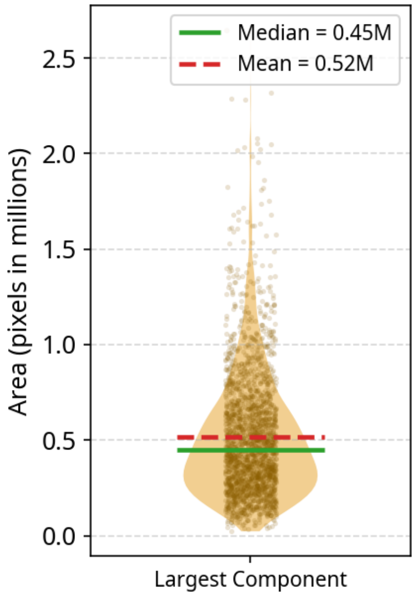


Figure S1: The fraction of area covered by the largest component (C1) of predicted masks for multi-region images in the whole annotated set.

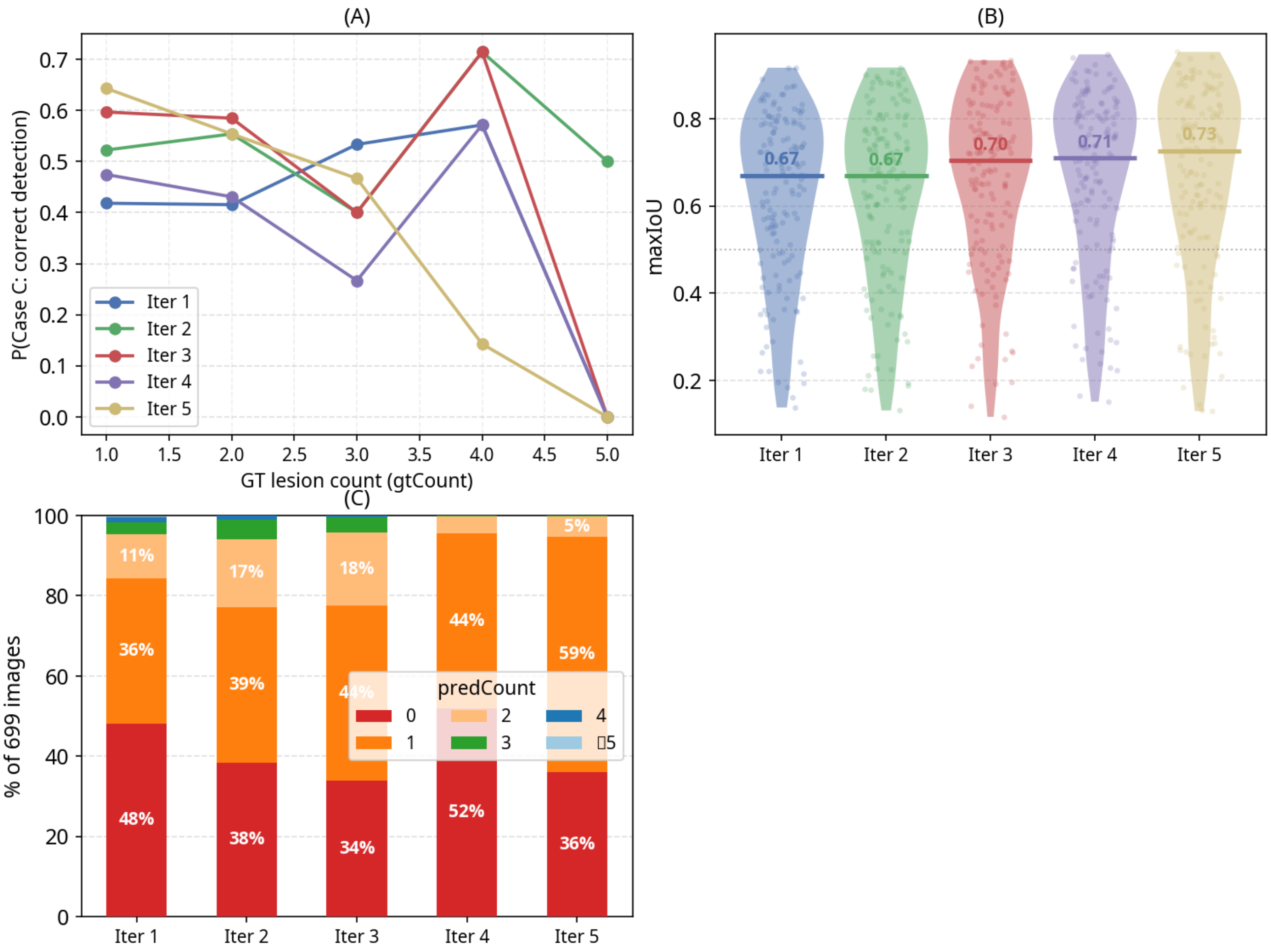


Figure S2: *Panel* (A) Relation between correct detection (Case C) and GT lesion (mask) count across all five iterations.(B) maxIoU distribution for the consistently detected cohort ($n = 145$, Case C in all 5 iters) across iterations; median rises from $0.67$ (iter. 1) to $0.73$ (iter. 5). (C) depicts the change in proportion of images with different numbers of predicted masks (0 to $\geq 5$).

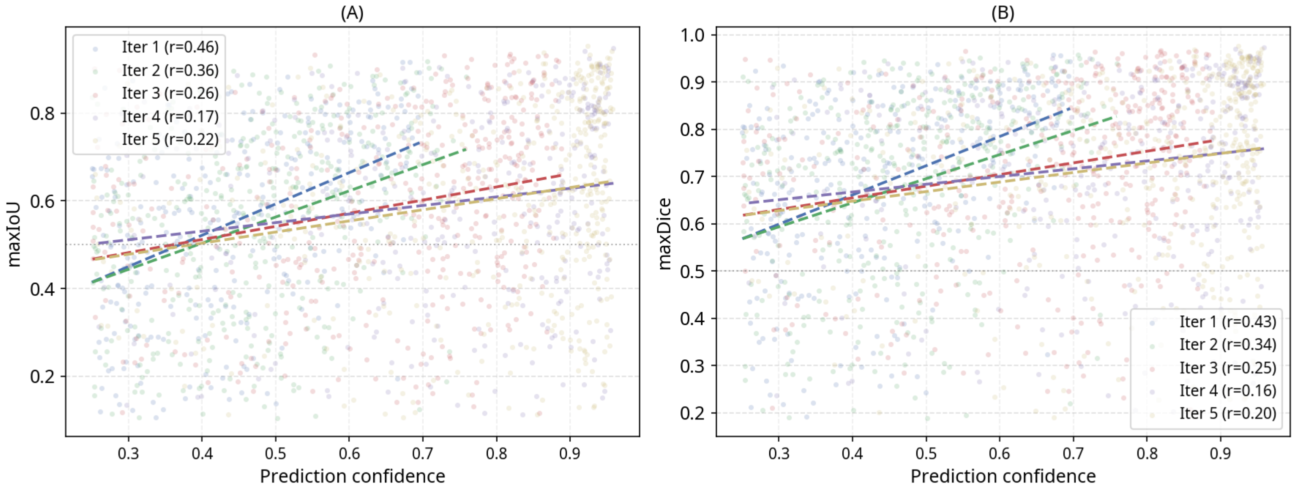


Figure S3: Scatterplots of `yoloConfidence1` vs. (A) `maxIoU` and (B) `maxDice`; dashed line = OLS trend; Pearson $r$ per iteration shown, Case C images only).
